# Stratification of individuals without prior diagnosis of diabetes using continuous glucose monitoring

**DOI:** 10.1101/2025.02.25.25322890

**Authors:** Hikaru Sugimoto, Gal Sapir, Ayya Keshet, Shinya Kuroda

**Affiliations:** Department of Biochemistry and Molecular Biology, Graduate School of Medicine, The University of Tokyo, 7-3-1 Hongo, Bunkyo-ku, Tokyo 113-0033, Japan; Department of Computer Science and Applied Mathematics, Weizmann Institute of Science, Rehovot, Israel; Pheno.AI, Tel-Aviv, Israel; Department of Molecular Cell Biology, Weizmann Institute of Science, Rehovot, Israel; Department of Biological Sciences, Graduate School of Science, The University of Tokyo, 7-3-1 Hongo, Bunkyo-ku, Tokyo 113-0033, Japan

## Abstract

An unmet need for preventing diabetes complications is the early detection of metabolic dysregulation. While glucose dynamics provide high-dimensional insights into metabolic states, extracting comprehensive and interpretable information from such data remains challenging. Here we show that the majority of inter-individual variation in glucose dynamics can be captured by just three features - mean, variance and autocorrelation - each independently associated with diabetes-related measures, even in individuals without a prior diabetes diagnosis. Analysis of continuous glucose monitoring data from 8,025 individuals showed that these three measures explained over 80% of the inter-individual variation in glucose dynamics. These measures outperformed conventional measures, including fasting, mean, and 2-hour postprandial glucose levels, in reconstructing postprandial glucose dynamics. Each feature showed independent associations with vascular or hepatic status. By condensing high-dimensional glucose dynamics into three interpretable features with minimal loss of information, this framework provides a basis for a more accurate diabetes risk assessment.

## INTRODUCTION

Diabetes is a major global health burden, affecting over 500 million people worldwide^1,2^. Beyond those with a formal diagnosis, many individuals have impaired glucose tolerance or remain undiagnosed^3^. Moreover, recent research suggests that traditional diagnostic tests – such as HbA1c, fasting blood glucose (FBG) and 2-hour postprandial glucose (PG120) – may fail to detect early changes in glucose dynamics that contribute to diabetic complications^4–7^, highlighting the urgent need for more refined screening methods.

Continuous glucose monitoring (CGM) has revealed substantial heterogeneity in glucose fluctuation patterns, even among individuals without a diabetes diagnosis^6,8,9^. These fluctuation patterns, including variability and autocorrelation, are associated with diabetes-related conditions such as atherosclerosis, hepatic steatosis and insulin resistance^6,7,10,11^.

However, several challenges have limited the clinical translation of these findings. First, the redundancy of many CGM-derived measures necessitates the identification of a minimal set of parameters that effectively capture inter-individual variation in glucose dynamics^8^.

Second, reference ranges for these measures remain poorly characterised in individuals without diagnosed diabetes^13^. Third, questions remain regarding the optimal time window for assessment, with emerging evidence suggesting that 1-hour postprandial glucose levels may provide valuable information beyond PG120^14^. Fourth, although CGM with machine learning approaches is promising, its clinical utility is limited by poor interpretability, overfitting, lack of standardised benchmarks or insufficient methodological transparency^6^.

Here, we aim to address these challenges by investigating whether the majority of inter-individual variability in CGM-derived measures and postprandial glucose dynamics can be explained by three measures: mean, variance and an autocorrelation-derived index (AC_Var)^6^. We further investigate whether these three measures have independent associations with diabetes-related parameters even in individuals without diabetes, and define reference ranges for these measures. Finally, we provide open-source tools for calculating these measures to facilitate future studies.

## RESULTS

### Glucose mean, variance and autocorrelation explain most of the inter-individual variability in glucose dynamics

To identify a minimal set of measures characterising inter-individual variability in glucose dynamics, we applied factor analysis to CGM-derived measures from 8,025 non-diabetic individuals (**Fig. 1a, Methods**). The minimum average partial analysis indicated that the CGM-derived measures had three factors, which together explained 82% of the total variance. The first three factors had strong loadings from variance-related measures (*e.g.*, CGM_Std and MAGE), mean-related measures (*e.g.*, CGM_Mean and CGM_Median), and autocorrelation-related measures (*e.g.*, AC_Mean and AC_Var), respectively (**Fig. 1b**).

**Fig. 1.**
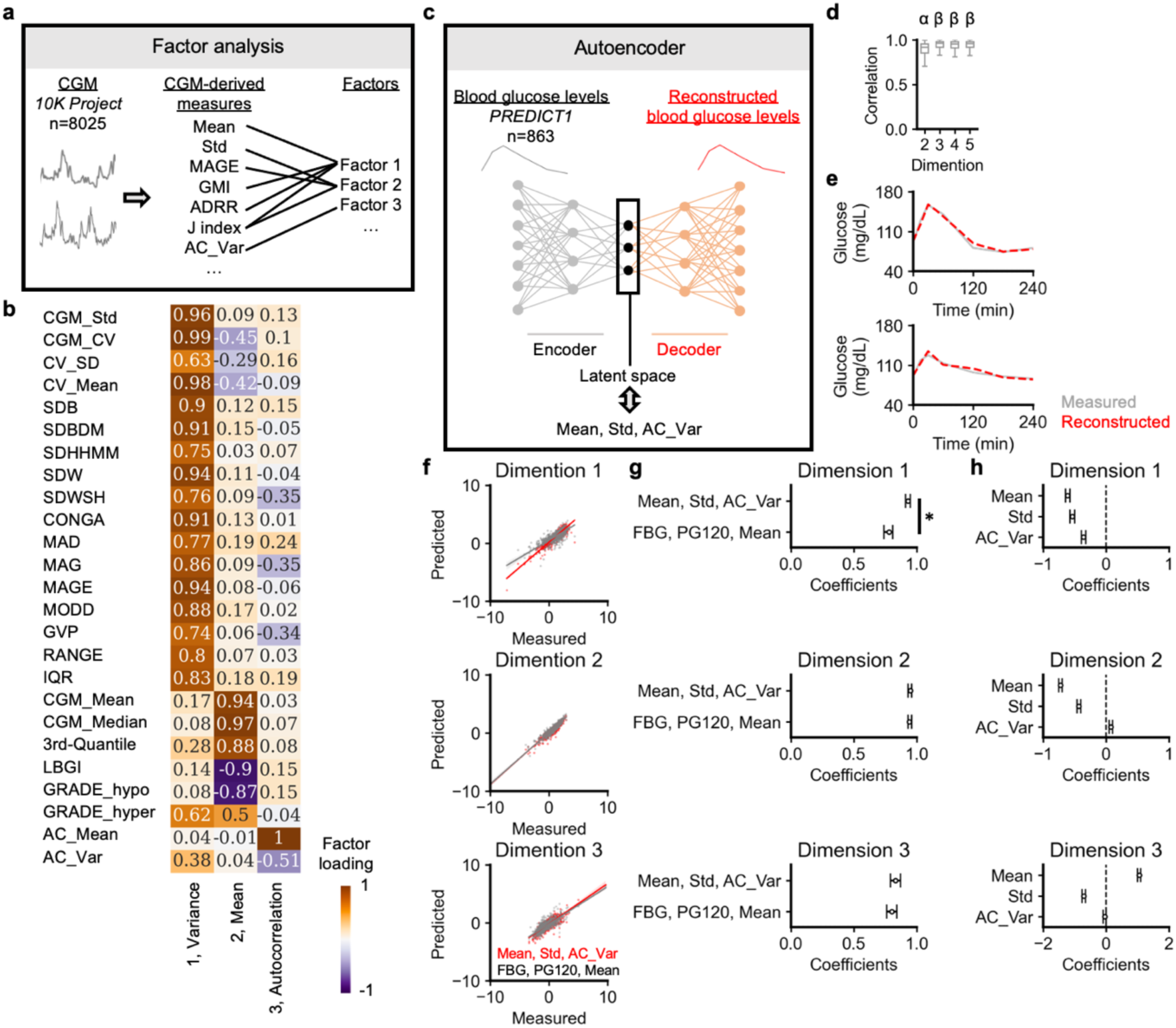
Decomposing glucose dynamics into key components. a, Schematic representation of the factor analysis approach applied to CGM data from the *10K Project* participants (**Methods**). **b,** Heatmap of factor loadings, showing the contributions of each CGM-derived index (rows) to the identified factors (columns). **c,** Schematic of performing autoencoder analysis on glucose dynamics during the meal challenge in the *PREDICT1 Study*. Latent space representations of glucose dynamics were compared with mean (Mean), standard deviation (Std) and autocorrelation (AC_Var) of glucose levels. **d,** Correlation between measured and reconstructed glucose levels across different latent space dimensions. Groups labelled with different letters are significantly different (*Q* < 0.05). **e,** Examples of measured and reconstructed glucose levels using three latent space dimensions. **f,** Comparison of measured and predicted values for each latent dimension using linear regression with Mean, Std, AC_Var as predictors (red). Grey shows predictions using FBG, PG120 and Mean. **g,** 95% confidence intervals for correlation coefficients between measured and predicted values using either Mean, Std, AC_Var (top) or FBG, PG120, Mean (bottom) as predictors. **h,** 95% confidence intervals for regression coefficients estimating the contribution of Mean, Std and AC_Var to the latent space dimensions. \**Q* < 0.05.

Similarly, principal component analysis revealed that first three principal components were associated with the mean, variance, or autocorrelation-related measures, together explaining 85% of the total variance (**Extended Data Fig. 1a, b**), suggesting that CGM-derived measures are mainly summarised by mean, variance, and autocorrelation.

To investigate whether postprandial glucose trajectories following standardised meals could also be summarised by these three features, we applied autoencoder models to glucose trajectory data from 863 non-diabetic individuals (**Fig. 1c, Methods**). A two-dimensional encoding achieved moderate reconstruction accuracy (R = 0.88, 95% CI: 0.87-0.88), while three dimensions significantly improved performance (R = 0.92, 95% CI: 0.92-0.93) (**Fig. 1d, e**). Additional dimensions provided minimal improvements (ΔR < 0.01), leading us to focus on the three-dimensional representation in subsequent analyses. We then examined how these latent dimensions related to the mean (Mean), standard deviation (Std), and autocorrelation (AC_Var) of glucose levels. Multiple regression analysis showed that Mean, Std and AC_Var accurately and independently predicted all three latent dimensions (R = 0.93, 95% CI: 0.91-0.95, R = 0.95, 95% CI: 0.93-0.96, and R = 0.83, 95% CI: 0.80-0.87) (**Fig. 1f-h**). These models outperformed those using FBG, PG120 and Mean, especially for the first latent dimension (*Q* < 0.05). Random forest models with Boruta confirmed the independent association of Mean, Std and AC_Var with the latent dimensions (**Extended Data Fig. 1c-e**). Multiple regression analysis followed by principal component analysis of the glucose trajectories showed independent associations between the three measures and principal components of glucose dynamics (**Extended Data Fig. 1f, g**). These results indicate that the combination of Mean, Std and AC_Var can accurately reconstruct the whole glucose trajectory profiles, whereas FBG, PG120 and Mean are insufficient for reconstruction.

To validate these results, we applied the autoencoder models to glucose fluctuations during 75 g oral glucose tolerance tests in an independent dataset (**Extended Data Fig. 2a**). The three-latent dimension model accurately reconstructed the glucose trajectories (R = 0.91, 95% CI: 0.89-0.93) (**Extended Data Fig. 2b, c**). The 2, 4, and 5 latent dimension models tended to be less accurate (R = 0.88, 95% CI: 0.86-0.90, R = 0.90, 95% CI: 0.88-0.92, R = 0.90, 95% CI: 0.88-0.92). Multiple regression analyses showed that Mean, Std, and AC_Var accurately and independently predicted all latent dimensions (R = 0.98, 95% CI: 0.97-0.99, R = 0.96, 95% CI: 0.94-0.98, and R = 0.90, 95% CI: 0.81-0.95) (**Extended Data Fig. 2d-f**). These prediction models performed better than those including FBG, PG120 and Mean, especially for the first latent dimension (*Q* < 0.05). Mean, Std, and AC_Var were independently associated with the disposition index, a marker of β cell function relative to insulin resistance (**Extended Data Fig. 2g-i**). These results validate that the combination of Mean, Std and AC_Var can reconstruct the whole glucose trajectory profile.

### Glucose mean, variance and autocorrelation are independently associated with vascular and hepatic health

We then examined the associations between the three glucose dynamics features and diabetes-related measures, including carotid intima-media thickness (IMT) and liver ultrasound-derived measures (attenuation, elasticity and dispersion) in 1,784 non-diabetic individuals (**Fig. 2a, Methods)**. IMT is a marker of atherosclerosis. Attenuation, elasticity and dispersion reflect hepatic fat content, stiffness and inflammation, respectively^16^.

**Fig. 2.**
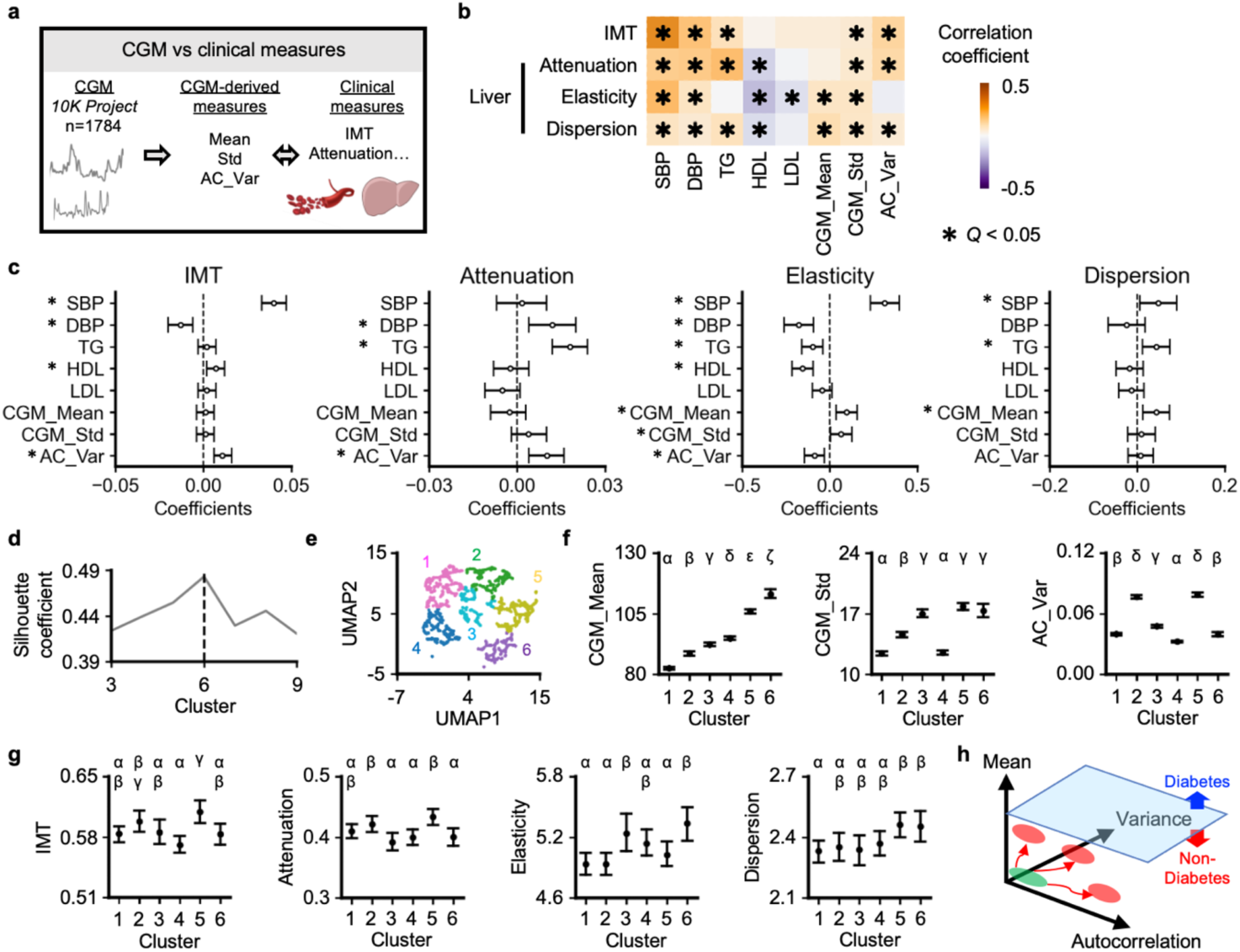
Clinical measures correlated with the three components of glucose dynamics. a, Schematic representation of analysing the relationships between CGM-derived measures (Mean, Std, AC_Var) and clinical measures, including IMT and liver ultrasound-based measures (attenuation, elasticity and dispersion). Data were obtained from the *10K Project* (**Methods**). **b,** Heatmap of Spearman correlation coefficients between CGM-derived measures and clinical parameters. \**Q* < 0.05. **c,** Multiple regression analysis showing the association between CGM-derived measures and the clinical parameters. Bars represent 95% confidence intervals of the regression coefficients. **d,** Silhouette coefficient as a function of the number of clusters, with the dotted line indicating the optimal number of clusters. **e,** UMAP of participant clustering based on CGM-derived measures (Mean, Std, AC_Var). **f,** Distributions of CGM-derived measures across clusters, presented as 95% confidence intervals. **g,** Cluster-wise distributions of the clinical parameters, presented as 95% confidence intervals. Groups labeled with different letters are significantly different (*Q* < 0.05). **h,** Graphical summary of inter-individual differences in glucose dynamics. Red indicates individuals with elevated diabetes-related clinical measures compared to those in green.

CGM_Mean, CGM_Std, and AC_Var were significantly correlated with IMT, attenuation, elasticity, or dispersion (*Q* < 0.05) (**Fig. 2b)**. Multiple regression analysis showed that AC_Var was significantly associated with IMT (β = 0.011, 95% CI: 0.006-0.016) and attenuation (β = 0.010, 95% CI: 0.004-0.016), even after adjustment for blood pressure (SBP and DBP), lipid profiles (TG, LDL-C, and HDL-C), CGM_Mean, and CGM_Std (**Fig. 2c**). Bayesian information criterion values of the models predicting IMT, attenuation, elasticity, and dispersion using the three measures were-3114,-2565, 5658, and 3170, respectively, which were lower than those obtained using models including 23 CGM-derived measures calculated with *iglu* package^17^ and shown in **Figure 1b** (-2886,-2373, 5868, and 3416, respectively), suggesting that the three measures accurately predict these parameters with fewer parameters. Moreover, CGM_Mean and AC_Var were independently associated with IMT even two years after CGM measurement (**Extended Data Fig. 3a-c**).

To establish reference ranges for these three measures, we performed a clustering analysis (**Fig. 2d, e**). Silhouette coefficients indicated that the optimal number of clusters was 6.

Cluster 2, characterised by high AC_Var but low CGM_Mean and CGM_Std, showed significantly higher liver attenuation compared to cluster 3 (*Q* < 0.05) (**Fig. 2f, g**). In contrast, cluster 6, characterised by high CGM_Mean and CGM_Std but low AC_Var, showed increased elasticity and dispersion. In two independent studies, individuals whose CGM-derived measures exceeded the lower 95% CI limits for high-value clusters (CGM_Mean: 111 mg/dL in cluster 6, CGM_Std: 17 mg/dL in cluster 3, AC_Var: 0.075 in cluster 2) had significantly lower insulin sensitivity or disposition index (*P* < 0.05 for both; **Extended Data Fig. 3d, e, Methods**), confirming the finding that CGM_Mean, CGM_Std, and AC_Var are associated with diabetes-related clinical measures even in individuals without a prior diagnosis of diabetes.

## DISCUSSION

This study shows that three measures (mean, variance, and autocorrelation) explain the majority of the inter-individual variability in glucose dynamics, with all other glucose dynamics-derived features derivable from these measures. In contrast, FBG and PG120 are insufficient to fully characterise this inter-individual variability, as they are primarily correlated only with mean glucose levels (**Extended Data Fig. 4a**). Notably, measures derived from mean, variance and autocorrelation showed independent associations with liver status and IMT. Collectively, these results indicate that while inter-individual variation exists across all three parameters, current diabetes diagnosis primarily relies on mean values (**Fig. 2h**, blue surface). Furthermore, even in non-diabetic individuals, elevated levels of any single parameter are associated with higher diabetes-related measures (**Fig. 2h**, red circles). CGM-based assessment of these three measures would provide a simpler and more accurate estimate of individual metabolic status than conventional diagnostic methods.

Beyond clinical applications, these findings open new research directions. Molecular studies, including genome-wide association analyses, have traditionally focused on mean or variance-related measures. The identification of autocorrelation as an independent component suggests the existence of previously unexplored physiological mechanisms that warrant further investigation. Furthermore, by demonstrating that the three measures can reconstruct entire glucose dynamics, our findings address key challenges in applying machine learning to glucose dynamics, including poor interpretability, low reproducibility, and overfitting due to the high dimensionality of glucose-derived features. This interpretable, simple and open-source framework provides a benchmark for future computational studies.

The current study has several limitations. Although statistically significant, the observed correlations were modest, likely due to the study population consisting mainly of healthy individuals with minimal inter-individual variability. Including individuals with impaired glucose tolerance may yield stronger associations. Furthermore, it remains to be determined whether CGM-based identification of at-risk individuals, followed by targeted interventions, improves outcomes.

In conclusion, inter-individual differences in glucose dynamics are primarily characterised by mean, variance and autocorrelation. Even in individuals without a prior diabetes diagnosis, these three features were independently associated with markers of vascular and hepatic health. These findings have the potential to refine screening and risk stratification for impaired glucose homeostasis, while also paving the way for further research into the physiological mechanisms underlying autocorrelation, a previously underappreciated glycaemic pattern. To make this advance easily accessible, we have developed a web application (https://cgmacapp.streamlit.app/) and a Python library (https://pypi.org/project/cgmac/) to calculate these measures.

## METHODS

### Study design and ethical approval

This study involves exploratory analyses of five independent observational studies that primarily examined glucose dynamics in individuals without a prior diabetes diagnosis^6,8,9,17–19^. In this paper, these studies were referred to as the *10K Project*^8,17^, the *PREDICT1 Study*^9^, the *San Francisco Bay Area Study1*^18^, the *San Francisco Bay Area Study2*^19^ and the *Kobe University Study*^6^. All studies were conducted in accordance with the Declaration of Helsinki and received approval from their respective ethics committees.

The *10K Project* is an ongoing observational prospective cohort study investigating glucose fluctuations using continuous glucose monitoring (CGM) in non-diabetic individuals^8,17^. The primary objective of this study was to assess interrelationships among CGM-derived measures, while the secondary objective was to assess the associations between CGM-derived measures and carotid/liver ultrasound derived-measures. The study was approved by the Institutional Review Board of the Weizmann Institute of Science. The data were obtained from https://humanphenotypeproject.org/home.

The *PREDICT1 Study* is an observational study investigating postprandial glucose responses to mixed-nutrient diets containing 86 g of carbohydrate and 53 g of fat administered in a tightly controlled clinical setting^9^. The current study analysed glucose levels up to four hours after the mixed-nutrient dietary challenges, which can be accessed at https://www.nature.com/articles/s41591-020-0934-0. The primary objective of this study was to test whether glucose-derived measures - mean, variance and autocorrelation - could reconstruct postprandial glucose responses more effectively than conventional indices, such as fasting glucose, two-hour postprandial glucose and mean glucose. This study was approved by the Research Ethics Committee and Integrated Research Application System (IRAS 236407).

The *San Francisco Bay Area Study1* is an observational study investigating the relationship between glucose levels during oral glucose tolerance tests and glucose control abilities^6^. The primary objective was to test whether glucose-derived measures - mean, variance and autocorrelation - could reconstruct the glucose dynamics better than fasting glucose, two-hour glucose and mean glucose. The study was approved by the Stanford University School of Medicine Human Research Protection Office (Institutional Review Board #43883).

The *San Francisco Bay Area Study2* is an observational study investigating the relationship between CGM-derived measures and insulin resistance quantified by the steady-state plasma glucose (SSPG) test^20^. The primary objective of the analysis was to assess the relationship between the three CGM-derived measures (mean, standard deviation, and autocorrelation function of glucose levels) and insulin resistance. The study was approved by the Stanford Institutional Review Board (IRB 37141).

The *Kobe University Study* is an observational study investigating the associations between CGM-derived measures and glucose control ability^6^. The primary objective of the analysis was to assess the relationship between the three CGM-derived measures (mean, standard deviation, and autocorrelation function of glucose levels) and the product of insulin secretion and insulin sensitivity assessed by hyperglycaemic and hyperinsulinemic-euglycaemic clamp tests (clamp disposition index, Clamp DI). The study was approved by the Ethics Committee of Kobe University Hospital (approval number 1834).

### Study population and inclusion criteria

A detailed description of the inclusion and exclusion criteria, as well as the measures obtained can be found in the previous publications^6,8,9,17–19^. A summary of the main criteria is given below and in **Extended Data Table 1**.

The *10K Project* recruited non-diabetic individuals aged 40-70 years between January 2019 and May 2023. Participants with more than seven days of FreeStyle Libre Pro Flash CGM data were included in the analysis. Exclusion criteria included self-reported diabetes and use of diabetes-related medications. Of the 9,529 participants at baseline, 8,025 met the inclusion criteria.

The *PREDICT1 Study* recruited non-diabetic individuals aged 18-65 years between June 2018 and May 2019. Participants with incomplete blood glucose data during the dietary challenges were excluded. Other exclusion criteria included active inflammatory disease, cancer, gastrointestinal disorders, pregnancy, recent antibiotic use, capillary glucose level of >216 mg/dL, type 1 diabetes mellitus, use of medication for type 2 diabetes mellitus, clinically diagnosed depression, recent heart attack or stroke, vegan, eating disorders or unwillingness to consume foods that are part of the study. Of the 1,002 participants at baseline, 863 met the inclusion criteria.

The *San Francisco Bay Area Study1* included participants aged 30-70 years with a body mass index (BMI) of 23-40 kg/m^2^. Exclusion criteria included major organ diseases, prior diagnosis of diabetes, uncontrolled hypertension, malignancy, chronic inflammatory conditions, use of any medications known to alter blood glucose or insulin sensitivity, haematocrit < 30, creatinine above the normal range, and ALT levels more than twice the upper limit of normal. Individuals with missing values were excluded from the analysis. Of the 56 participants screened, 51 met the inclusion criteria. After screening tests, 8, 27, and 16 individuals met the criteria of type 2 diabetes mellitus, pre-diabetes, and normal glucose tolerance, respectively.

The *San Francisco Bay Area Study2* included participants aged 25-76 years with no prior diagnosis of diabetes who wore a Dexcom G4 CGM for more than three days and underwent SSPG testing. Exclusion criteria included major organ diseases, chronic inflammatory conditions, malignancy, uncontrolled hypertension, eating disorders, history of bariatric surgery, diagnosis of diabetes, use of weight-loss or diabetogenic medications, or recent unstable weight. Of the 57 participants screened, 41 met the inclusion criteria. After screening tests, 4, 9, and 28 individuals met the criteria of type 2 diabetes mellitus, pre-diabetes, and normal glucose tolerance, respectively.

The *Kobe University Study* recruited individuals aged 20-75 years with no prior diagnosis of diabetes between January 2016 and May 2018. Participants with more than three days of iPro CGM data were included. Exclusion criteria included medications affecting glucose metabolism, psychiatric disorders, and pregnancy. Of the 70 participants initially recruited, 64 met the inclusion criteria. After screening tests, 3, 9, and 52 individuals met the criteria of type 2 diabetes mellitus, impaired glucose tolerance, and normal glucose tolerance, respectively.

### Calculation of glucose dynamics-derived measures

CGM-derived measures were calculated using the *iglu* package^19^ and custom scripts (https://pypi.org/project/cgmac/), following the methodology of previous studies^6,8^. AC_Mean and AC_Var were calculated as the mean and variance of glucose autocorrelation functions over lags 1-10, using 15-minute interval data. To maintain consistency with the *10K Project*, CGM data collected at 5-minute intervals in the *San Francisco Bay Area Study2* and the *Kobe University Study* were resampled to 15-minute intervals. A brief description of the CGM-derived measures analysed is shown in **Extended Data Table 2**.

In the *PREDICT1 Study*, blood glucose levels were measured at 0, 15, 30, 60, 120, 180 and 240 minutes during the dietary challenge. In the *San Francisco Bay Area Study1*, blood glucose levels were measured at 0, 10, 15, 20, 30, 40, 50, 60, 75, 90, 105, 120, 135, 150 and 180 minutes during OGTT. Data were linearly interpolated to obtain 15-minute interval values for calculation of the mean, standard deviation and AC_Var. For validation of autoencoder models trained on the *PREDICT1 Study* data using data from the *San Francisco Bay Area Study1*, blood glucose variability beyond 180 minutes post-OGTT was assumed to be minimal, and blood glucose values at 180 and 240 minutes were set equal.

### Clinical measures and assessments

A detailed description of the clinical measures analysed in this study can be found in the previous publications^6,8,9,17,18^.

In the *10K Project*, ultrasound assessments of carotid intima-media thickness (IMT), liver attenuation, elasticity and dispersion were performed. Ultrasound examinations were performed in 1,784 participants at the same time as their CGM assessment, and in 1,023 participants two years later. Due to the lack of overlap between these groups and the inability to track longitudinal changes over the two-year period, the associations between the ultrasound-derived measures and the CGM-derived measures were analysed separately.

Systolic blood pressure (SBP), diastolic blood pressure (DBP), triglycerides (TG), low-density lipoprotein cholesterol (LDL-C), and high-density lipoprotein cholesterol (HDL-C) measured at the time of the CGM assessment were also included in the analyses.

In the *San Francisco Bay Area Study1*, β-cell function was quantified by calculating the insulin secretion rate using the C-peptide deconvolution method. Insulin resistance was quantified using the SSPG test. Individuals with greater insulin resistance would have higher SSPG concentrations. The disposition index was calculated as the area under the insulin secretion rate divided by SSPG.

In the *San Francisco Bay Area Study2*, insulin resistance was assessed using the SSPG test. After an overnight fast, participants received continuous intravenous infusions of octreotide (0.27 μg/m^2^/min), insulin (25 mU/m^2^/min) and glucose (240 mg/m^2^/min) for 180 minutes. Blood samples were taken at 10-minute intervals during the last 30 minutes of the infusion.

In the *Kobe University Study*, insulin secretion was quantified as the incremental area under the insulin curve during the first 10 minutes of a hyperglycaemic clamp test. Insulin sensitivity was calculated as the mean glucose infusion rate during the last 30 minutes of the hyperinsulinemic-euglycaemic clamp tests, normalised to plasma glucose and serum insulin concentrations. The disposition index (Clamp DI) was defined as the product of insulin secretion and sensitivity.

### Statistical analyses

Relationships among CGM-derived measures were assessed using exploratory factor analysis (EFA) and principal component analysis (PCA). EFA groups observed variables into a smaller set of unobserved, uncorrelated factors, while PCA identifies major axes of variation in high-dimensional data. Variables with factor loadings of ≥ 0.40 were retained for interpretation following Oblimin rotation. Prior to the analyses, all data were standardised using z-score normalisation.

Several CGM-derived measures, such as time above 250 mg/dL, were excluded from the analyses because their values were close to zero in most non-diabetic individuals (**Extended Data Fig. 4b**). These non-normally distributed measures (**Extended Data Fig. 4c**), highly correlated measures (correlations ≥ 0.99, *e.g.*, mean glucose and glucose AUC), and composite measures such as the J-index (0.001×(CGM_Mean+CGM_Std)^2^) were excluded. These exclusions were applied to reduce redundancy and avoid singular covariance matrices issues in EFA (for a discussion on singular covariance matrices, see https://stats.stackexchange.com/questions/11102/factor-analysis-problem-singular-covariance-matrix). While PCA is more robust to singular covariance matrices and allows the inclusion of highly correlated and composite variables, EFA requires a more selective set of variables. The final analyses included 25 variables for EFA (**Fig. 1b**) and 34 variables for PCA (**Extended Data Fig. 1b**). These analyses were performed using factor-analyzer 0.5.1 (https://pypi.org/project/factor-analyzer/) and scikit-learn 1.0.2 (https://scikit-learn.org/stable/).

Prior work has categorized CGM-derived measures into five groups: “mean”, “variability”, “in range”, “high glucose”, and “low glucose”, using Pearson correlation and hierarchical clustering^8^. While these methods provide valuable insights into the characteristics of CGM-derived measures, they can lead to over-classification due to methodological limitations. For example, composite variables often have weaker individual correlations with primary variables, despite being derivable from their combinations. Indeed, measures categorized under “high glucose” and “low glucose”, such as time above 180 mg/dL and time below 70 mg/dL, were significantly associated with both “mean” and “variability” (**Extended Data Fig. 4d**). Furthermore, the “in range” category is mathematically dependent on “high glucose” and “low glucose” (*e.g.*, time in range 70-180 mg/dL can be predicted from time above 180 mg/dL and time below 70 mg/dL). To address these limitations, this study applied EFA and PCA to identify a minimal yet comprehensive set of CGM-derived measures that effectively capture glucose dynamics. Additionally, recently developed autocorrelation-based measures, such as AC_Mean and AC_Var, were included, as they have been shown to correlate significantly with insulin sensitivity and coronary artery lesions, independently of conventional CGM-derived measures such as mean and variance^6,7^.

Autoencoder models were implemented using TensorFlow (https://www.tensorflow.org) following the methodology of a previous study^20^. In this study, the autoencoder models were used to identify latent dimensions of postprandial glucose responses while preserving the ability to reconstruct the original glucose patterns. The encoder component consisted of sequential layers, starting with an input layer containing 17 nodes, followed by a dense layer of 12 nodes with LeakyReLU activation. This was followed by batch normalisation and dropout at a rate of 0.1. The next layer included a dense layer of 8 nodes with LeakyReLU activation, followed by batch normalisation. The encoder ended with a bottleneck layer of 3 nodes using linear activation and Glorot normal initialisation. The decoder architecture mirrored the encoder structure in reverse order, starting with a dense layer of 8 nodes with LeakyReLU activation, followed by batch normalisation and dropout. This was followed by a dense layer of 12 nodes with LeakyReLU activation and batch normalisation, and finally an output layer of 17 nodes. The network was trained using mean squared error loss and the Adam optimiser for 500 epochs with a batch size of 32. Twenty percent of the data was reserved for validation during training.

To evaluate the effect of different latent space dimensions, additional models with bottleneck layers of 2, 4 and 5 nodes were trained for comparison. Reconstruction performance of these models was compared using Pearson’s correlation coefficients between measured and reconstructed glucose levels. Initially, the model was developed using glucose time series data from the *PREDICT1 Study*. To assess its generalisability under entirely different conditions, the trained model was subsequently tested on glucose time series data from the *San Francisco Bay Area Study1*.

After training, 3-dimensional compressed representations of the glucose dynamics were extracted using the encoder part of the network. Relationships between these compressed features and three measures derived from glucose dynamics (mean glucose, glucose standard deviation and AC_Var) were analysed using linear regression and random forest models.

When running random forest models, hyperparameter optimisation was performed using a grid search with 10-fold cross-validation. The hyperparameter grid explored different values for maximum tree depth (10, 20, 30), minimum samples required for splitting (5, 10, 15) and minimum samples per leaf node (2, 4, 8). The number of trees was fixed at 300. Model performance was assessed using root mean squared error. Feature importance was assessed using random forest feature importance scores and the Boruta algorithm (https://github.com/scikit-learn-contrib/boruta_py).

In this study, AC_Var was chosen over AC_Mean based on the a priori hypothesis that AC_Var would characterise glucose dynamics more effectively than AC_Mean. This decision was supported by previous findings demonstrating the superior robustness of AC_Var to noise compared to AC_Mean^6^. In addition, AC_Var showed a significant correlation with AC_Mean (R = –0.64, 95% CI: –0.66 to –0.63) in the *10K Project* dataset. Previous studies have shown that CGM_Mean, CGM_Std, and AC_Var are independently associated with coronary artery disease and insulin sensitivity^6,7^. Consistent with these findings, factor analysis of the CGM-derived measures in this study identified three distinct factors, each of which included one of these three measures. Based on this evidence, the investigation focused on the relationships between CGM_Mean, CGM_Std, and AC_Var and clinical outcomes.

Group comparisons were performed using Welch’s t-test. Associations between variables were assessed using Spearman’s correlation coefficients and linear regression models.

Multiple comparisons were adjusted using the Benjamini-Hochberg procedure, with statistical significance set at *Q* < 0.05. To assess the predictive performance of models including different sets of CGM-derived measures, the Bayesian Information Criterion (BIC) was calculated for each model. Bootstrap resampling with 10,000 iterations was used to estimate 95% confidence intervals. These analyses were performed using statsmodels 0.13.2 (https://www.statsmodels.org/stable/index.html).

K-Means clustering was applied using CGM_Mean, CGM_Std and AC_Var as input features. Data were z-score normalised prior to clustering. Cluster quality was assessed using silhouette analysis. Results were visualised using Uniform Manifold Approximation and Projection (UMAP). These analyses were performed using scikit-learn 1.0.2 (https://scikit-learn.org/stable/) and umap-learn 0.5.7 (https://pypi.org/project/umap-learn/).

## Data availability

The data supporting the findings of this study originate from the *10K Project*^8,17^, the *PREDICT1 Study*^9^, the *San Francisco Bay Area Study1*^19^, the *San Francisco Bay Area Study2*^18^, and the *Kobe University Study*^6^. Data from the *10K Project* are available at https://humanphenotypeproject.org/. Data from the *PREDICT1 Study* are managed by the Department of Twin Research at Kings College London and can be accessed by researchers using the procedures described at https://twinsuk.ac.uk/ resources-for-researchers/access-our-data/. We only used data from https://www.nature.com/articles/s41591-020-0934-0. Data from the *San Francisco Bay Area Study1*^19^ (https://www.nature.com/articles/s41551-024-01311-6), the *San Francisco Bay Area Study2*^18^ (https://journals.plos.org/plosbiology/article?id=10.1371/journal.pbio.2005143), and the *Kobe University Study*^6^ (https://www.medrxiv.org/content/10.1101/2023.09.18.23295711v1) can be obtained from the previous publications^6,18,19^.

## Code availability

The web-based server calculating CGM-derived measures (https://cgmacapp.streamlit.app/) is implemented in Python and the Streamlit library (https://www.streamlit.io). A stand-alone tool to run the web application on a local machine is available at the GitHub repository (https://github.com/HikaruSugimoto/cgmac_app). The Python library calculating CGM-derived measures is available from https://pypi.org/project/cgmac/.

## Author contributions

H.S. conceived the project, designed and conducted the analyses, interpreted the results and wrote the manuscript. G.S. and A.K. curated the database. S.K. reviewed the paper and supervised the study.

## Funding and Assistance

This study was supported by the Japan Society for the Promotion of Science (JSPS) KAKENHI (JP21H04759), CREST, the Japan Science and Technology Agency (JST) (JPMJCR2123), The Uehara Memorial Foundation and The Takeda Science Foundation.

## Data Availability

All data produced are available online at
https://journals.plos.org/plosbiology/article?id=10.1371/journal.pbio.2005143
https://www.nature.com/articles/s41551-024-01311-6
https://humanphenotypeproject.org/
https://www.nature.com/articles/s41591-020-0934-0
https://www.medrxiv.org/content/10.1101/2023.09.18.23295711v1

---

**Extended Data Fig. 1.**
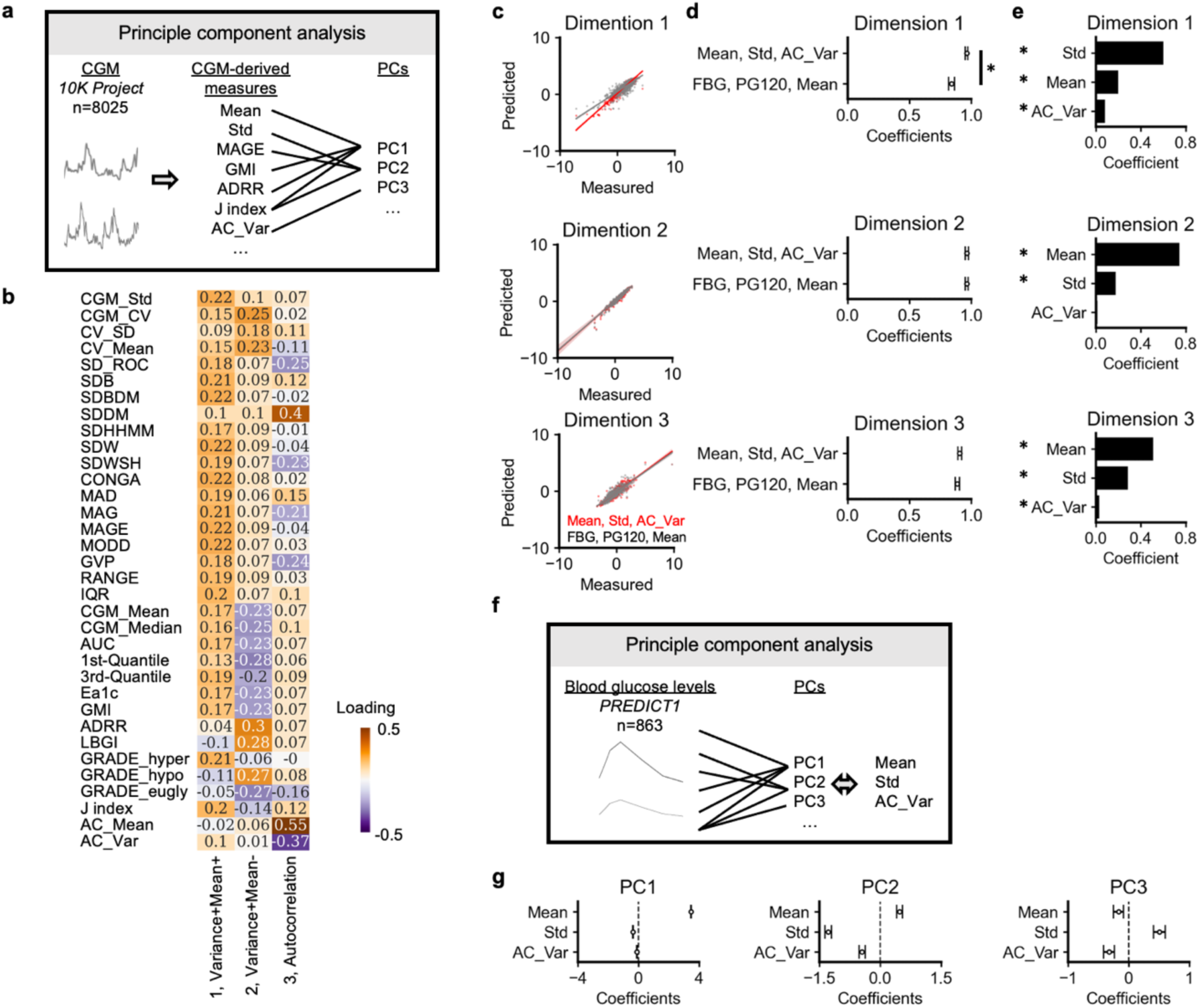
Principal component and factor analysis of glucose dynamics. a, Schematic representation of principal component analysis (PCA) applied to CGM data from the *JOK Project* participants **(Methods). b,** Heatmap of PCA loadings showing the contribution of each CGM-derived index (rows) to the principal components (columns). Colours represent the magnitude and direction of the loadings. **c,** Comparison of measured and predicted values for each latent dimension using random forest models. Red represents predictions using Mean, Std and AC_Var as predictors, while grey represents predictions using fasting blood glucose (FBG), 120-minute plasma glucose (PG120) and Mean. **d,** 95% confidence intervals for correlation coefficients between measured and predicted values across different sets of predictors. **e,** Feature importance scores from random forest models indicating the contribution ofCGM-derived measures to predictions. Statistical significance was assessed using the Boruta algorithm.*\*Q* < 0.05. **f,** Schematic representation of PCA applied to blood glucose levels after breakfast meal challenges in the *PREDICT! Study.* Relationships between principal components and CGM-derived measures (Mean, Std, AC_Var) were examined. **g,** 95% confidence intervals for regression coefficients estimating the contributions of Mean, Std and AC_Var to the principal components.

**Extended Data Fig. 2.**
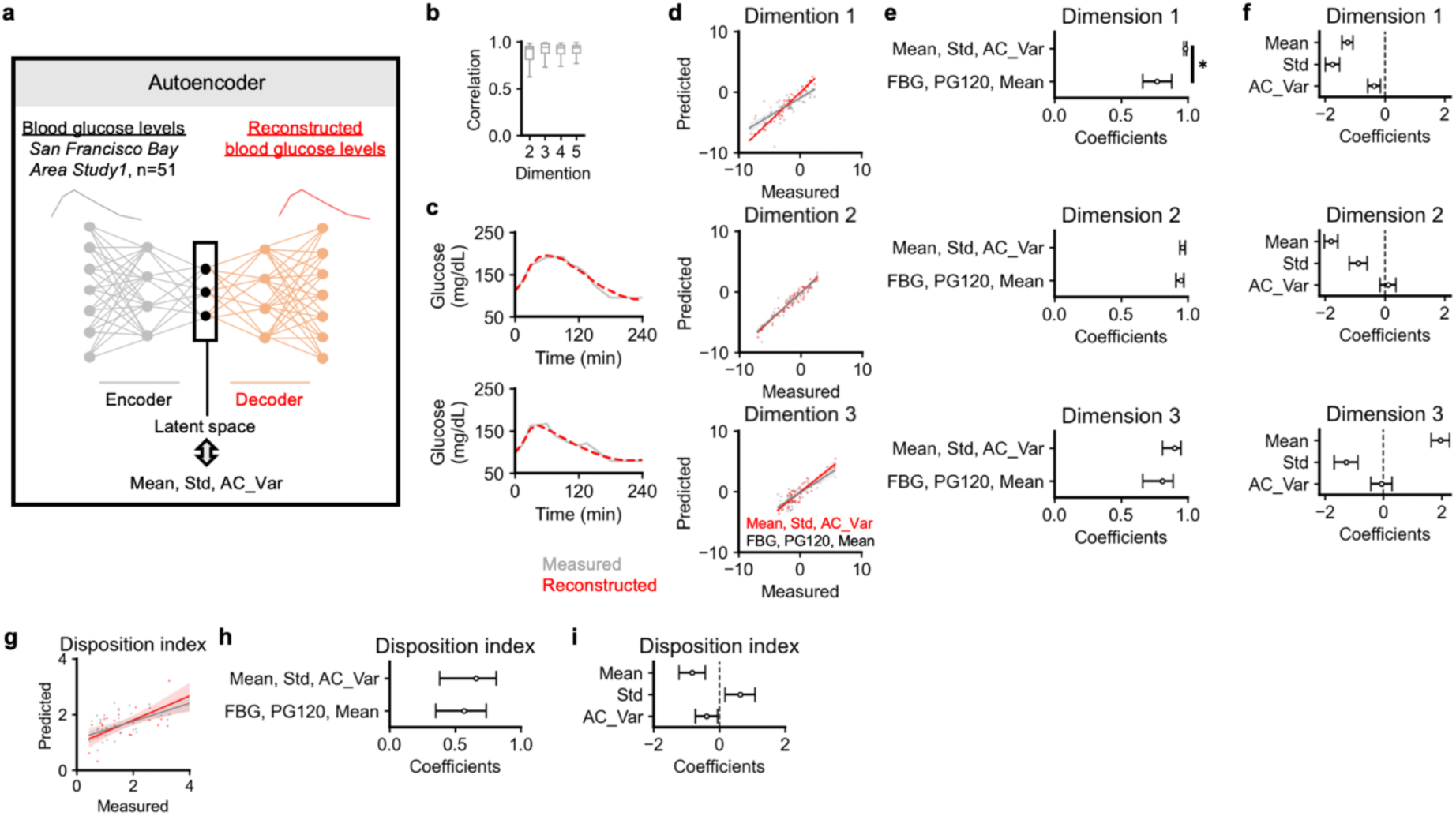
Latent dimensions of glucose dynamics during oral glucose tolerance tests. a, Schematic representation of performing autoencoder analysis on glucose dynamics during oral glucose tolerance tests performed in the *San Francisco Bay Area StudyI* **(Methods).** Latent space representations of glucose dynamics were compared with mean (Mean), standard deviation (Std) and autocorrelation (AC_Var) of glucose levels. **b,** Correlation between measured and reconstructed glucose levels across different latent space dimensions. **c,** Examples of measured and reconstructed glucose levels using three latent space dimensions. **d,** Comparison of measured and predicted values for each latent dimension using linear regression with Mean, Std, AC_Var as predictors (red). Grey shows predictions using FBG, PG120 and Mean. **e,** 95% confidence intervals for correlation coefficients between measured and predicted values using either Mean, Std, AC_Var (top) or FBG, PG120, Mean (bottom) as predictors. *\*Q* < 0.05. **f,** 95% confidence intervals for regression coefficients estimating the contribution of Mean, Std and AC_Var to the latent space dimensions. **g,** Comparison of measured and predicted disposition index using linear regression with Mean, Std, AC_Var as predictors (red). Grey shows predictions using FBG, PG120 and Mean. **h,** 95% confidence intervals for correlation coefficients between measured and predicted disposition index using either Mean, Std, AC_Var (top) or FBG, PG120, Mean (bottom) as predictors. i, 95% confidence intervals for regression coefficients estimating the contribution of Mean, Std and AC_Var to the disposition index.

**Extended Data Fig. 3.**
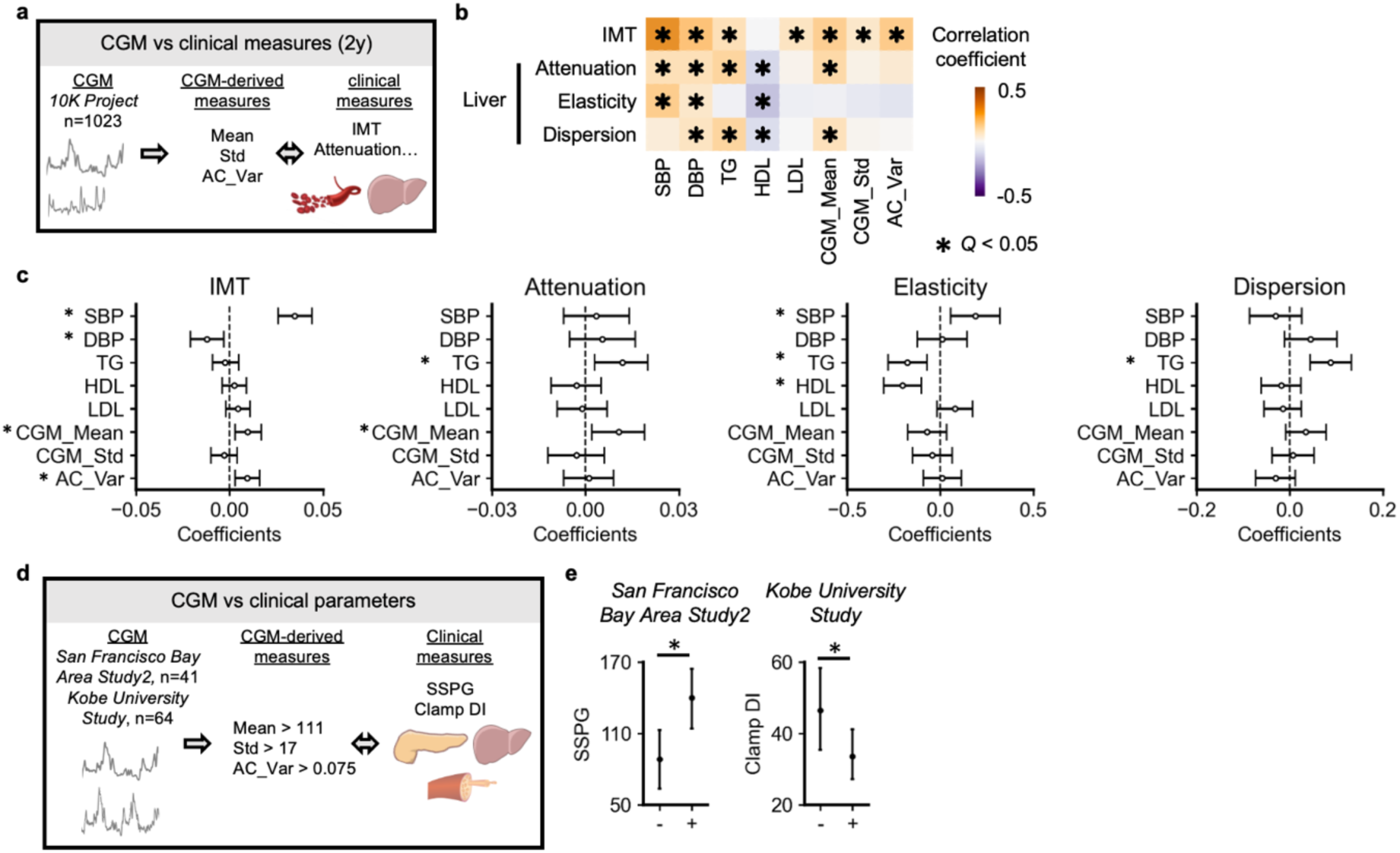
Associations between components of glucose dynamics and clinical measures. a, Schematic representation of analysing the relationships between CGM-derived measures (Mean, Std, AC_Var) and clinical measures, including intima-media thickness (IMT) and liver ultrasound-based measures such as attenuation, elasticity and dispersion measured two years after performing CGM. Data were obtained from the *JOK Project* **(Methods). b,** Heatrnap of Spearman correlation coefficients between CGM-derived measures and clinical parameters. *\*Q* < 0.05. c, Multiple regression analysis showing the association between CGM-derived measures (Mean, Std, AC_Var) and the clinical parameters. Bars represent 95% confidence intervals of the regression coefficients. **d,** Schematic representation of analysing the relationships between CGM-derived measures (Mean, Std, AC_Var) and markers of insulin resistance (steady-state plasma glucose, SSPG) and the product of insulin sensitivity and secretion capacity (clamp disposition index, Clamp DI). Higher SSPG indicates greater insulin resistance, whereas lower Clamp DI reflects reduced insulin sensitivity and secretion. Data were obtained from the *San Francisco Bay Area Study2* and the *Kobe University Study* **(Methods). e,** Bar plots ofSSPG and Clamp DI values stratified by CGM-derived measurement thresholds(+: Mean> 111 mg/dL, Std> 17 mg/dL, AC_Var> 0.075). Bars represent 95% confidence intervals. Welch’s t-test was used for group comparisons *(*P<* 0.05).

**Extended Data Fig. 4.**
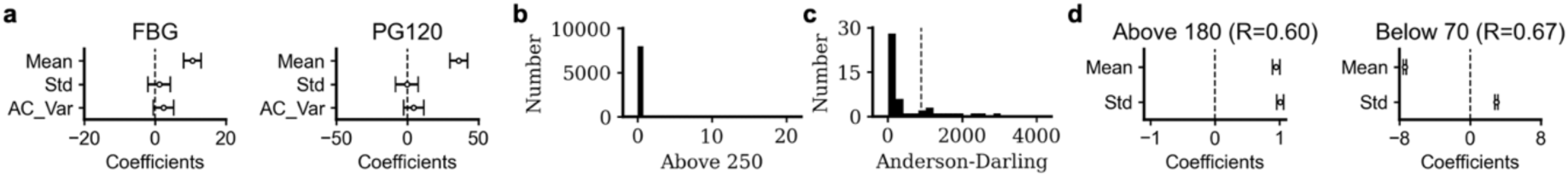
Characteristics of glucose dynamics-derived measures. a, Multiple regression analysis showing associations between glucose dynamics-derived measures (Mean, Std, AC_Var) and fasting blood glucose (FBG) and 2-hour plasma glucose (PGl20) levels during oral glucose tolerance tests. Data were from the *San Francisco Bay Area Study}.* Bars represent 95% confidence intervals of the regression coefficients. **b,** Distribution of time periods with glucose levels above 250 mg/dL in continuous glucose monitoring (CGM) data. Data were obtained from the *1OK Project.* **c,** Anderson-Darling test statistics for CGM­ derived measures in the *JOK Project.* Measures exceeding the threshold for non-normality (dotted line determined by the Otsu method) were excluded from subsequent analyses. **d,** Multiple regression analysis showing associations between glucose dynamics­ derived measures (Mean, Std) and time periods with glucose levels above 180 mg/dL or below 70 mg/dL. Data were obtained from the *1OK Project.* R is the correlation coefficient between measured and predicted values. Bars represent 95% confidence intervals of the regression coefficients.

**Extended Data Table 1.** Summary of participant characteristics. This study consists of exploratory analyses of the data from the *1OK Project,* the *PREDICT] Study,* the *San Francisco Bay Area Study],* the *San Francisco Bay Area Study2* and the *Kobe University Study.* Only summary statistics are presented due to differences in data sharing policies across studies, with some studies providing only aggregated data (e.g., age ranges rather than individual ages). Detailed participant selection criteria and measurement protocols for each study are described in the previous studies.

|  | *10K Project* | *PREDICT1 Study* | *San Francisco Bay Area Study1* | *San Francisco Bay Area Study2* | *Kobe University Study* |
| --- | --- | --- | --- | --- | --- |
| N | 8025 | 863 | 51 | 41 | 64 |
| Age | 40-70 | 18-65 | 30-70 | 25-76 | 20-75 |
| Diagnosis (Normal, Pre-DM, DM) | 8025, 0, 0 | 863, 0, 0 | 16, 27, 8 | 28, 9, 4 | 52, 9, 3 |

**Extended Data Table 2.**
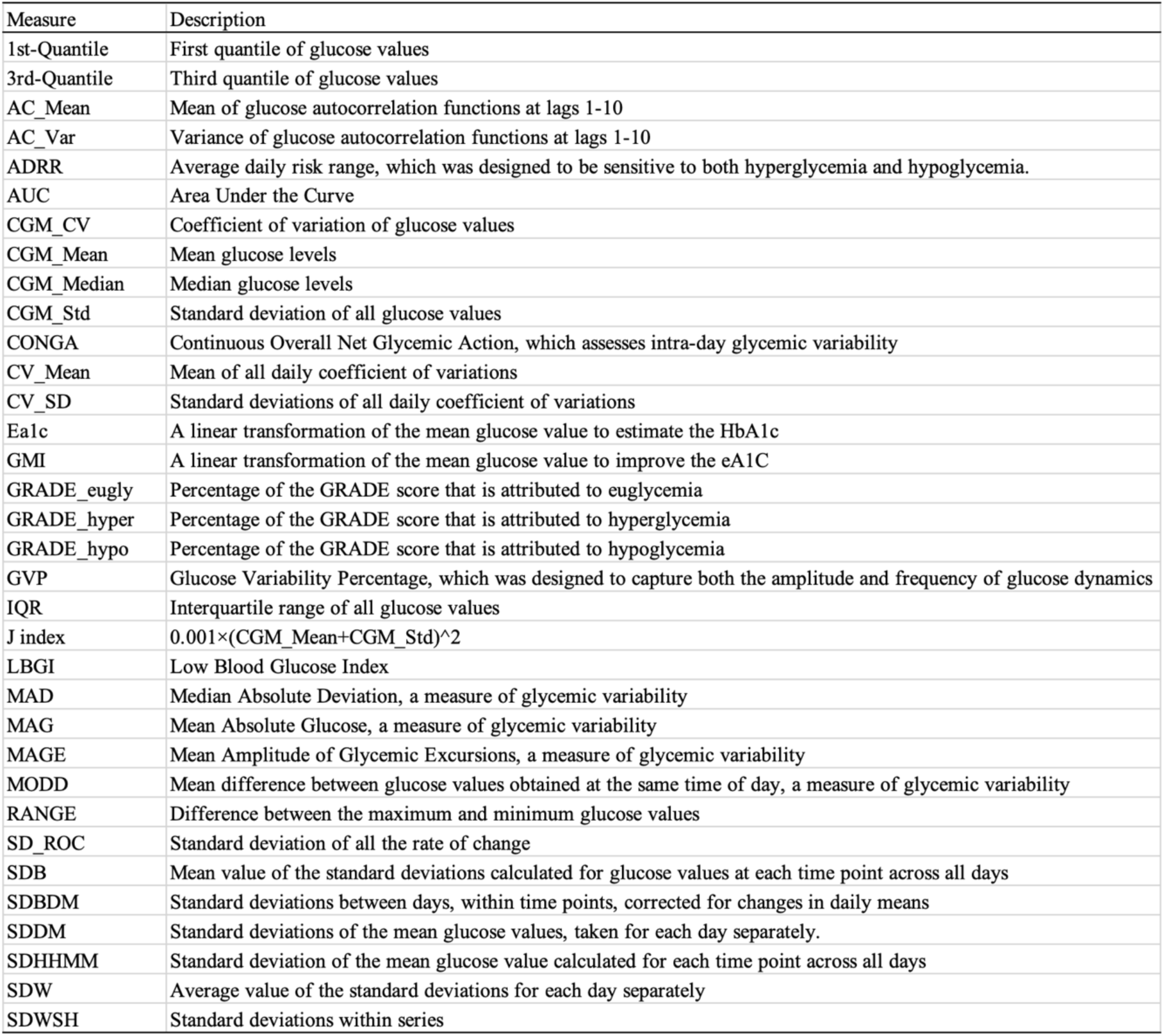
Summary of CGM-derived measures. A detailed description of each COM-derived measure can be found in previous studies.

| Measure | Description |
| --- | --- |
| 1st-Quantile | First quantile of glucose values |
| 3rd-Quantile | Third quantile of glucose values |
| AC_Mean | Mean of glucose autocorrelation functions at lags 1-10 |
| AC_Var | Variance of glucose autocorrelation functions at lags 1-10 |
| ADRR | Average daily risk range, which was designed to be sensitive to both hyperglycemia and hypoglycemia. |
| AUC | Area Under the Curve |
| CGM_CV | Coefficient of variation of glucose values |
| CGM_Mean | Mean glucose levels |
| CGM_Median | Median glucose levels |
| CGM_Std | Standard deviation of all glucose values |
| CONGA | Continuous Overall Net Glycemic Action, which assesses intra-day glycemic variability |
| CV_Mean | Mean of all daily coefficient of variations |
| CV_SD | Standard deviations of all daily coefficient of variations |
| Ea1c | A linear transformation of the mean glucose value to estimate the HbA1c |
| GMI | A linear transformation of the mean glucose value to improve the eA1C |
| GRADE_eugly | Percentage of the GRADE score that is attributed to euglycemia |
| GRADE_hyper | Percentage of the GRADE score that is attributed to hyperglycemia |
| GRADE_hypo | Percentage of the GRADE score that is attributed to hypoglycemia |
| GVP | Glucose Variability Percentage, which was designed to capture both the amplitude and frequency of glucose dynamics |
| IQR | Interquartile range of all glucose values |
| J index | $0.001 \times (\text{CGM\_Mean} + \text{CGM\_Std})^2$ |
| LBGI | Low Blood Glucose Index |
| MAD | Median Absolute Deviation, a measure of glycemic variability |
| MAG | Mean Absolute Glucose, a measure of glycemic variability |
| MAGE | Mean Amplitude of Glycemic Excursions, a measure of glycemic variability |
| MODD | Mean difference between glucose values obtained at the same time of day, a measure of glycemic variability |
| RANGE | Difference between the maximum and minimum glucose values |
| SD_ROC | Standard deviation of all the rate of change |
| SDB | Mean value of the standard deviations calculated for glucose values at each time point across all days |
| SDBDM | Standard deviations between days, within time points, corrected for changes in daily means |
| SDDM | Standard deviations of the mean glucose values, taken for each day separately. |
| SDHHMM | Standard deviation of the mean glucose value calculated for each time point across all days |
| SDW | Average value of the standard deviations for each day separately |
| SDWSH | Standard deviations within series |

